# Should WHO-prequalified antigen-antibody fourth-generation rapid diagnostic tests be used to detect acute HIV infection? A systematic review and meta-analysis of diagnostic performance

**DOI:** 10.64898/2026.03.19.26347008

**Authors:** Margot Neveux, Runa Hylin, Veronica Ruiz Gonzalez, Abril Patricia Lopez Parra, Ali Onoja, Sunee Sirivichayakul, Akanmu A Sulaimon, Gallican Rwibasira, Missiani Ochwoto

**Author notes:** **Corresponding Author:** Veronica Ruiz Gonzalez, Clinica Especializada Condesa, Mexico, Gral. Benjamín Hill 24, Hipódromo Condesa, Cuauhtémoc, 06170 Ciudad de México, CDMX, Mexico. **Email addresses of authors:** Margot Neveux, Runa Hylin, Veronica Ruiz Gonzalez, Abril P. Lopez Parra, Ali Onoja, Sunee Sirivichayakul, Akanmu Alani Sulaimon, Gallican Rwibasira, Missiani Ochwoto.

## Abstract

**Introduction:** Diagnostics have become the fundamental backbone of HIV prevention, treatment and long-term retention in care, and are critical to achieving the 95-95-95 UNAIDS targets. To effectively reach underserved and remote populations, diagnostic technologies must be cost-effective, robust, user-friendly and suitable for settings with limited infrastructure. Among available testing modalities, rapid diagnostic tests (RDTs) play a central role in expanding HIV testing coverage. Earlier generations of RDTs were limited by their inability to detect acute HIV, with limited ability to detect p24 antigen (Ag), an early marker of HIV infection, which is expected to shorten the diagnostic window to two-to-three weeks. The introduction of fourth-generation RDTs, which detects both chronic and acute HIV infection through p24 Ag detection, was designed to ensure that the traditional diagnostic window of two-to-three months is shortened to approximately two-to-three weeks. However, integrating these assays into existing testing algorithms requires clear evidence that they meet high standards of quality and performance. This systematic review aims to assess the performance of WHO-prequalified fourth-generation Ag/Ab RDTs.

**Methods:** We performed a systematic search across six databases to identify studies evaluating Ag/Ab RDTs against laboratory reference standards in individuals aged 12 years and older, spanning 1 January 2010 to 31 December 2025. Outcomes were limited to measures of diagnostic accuracy. A meta-analysis focusing exclusively on WHO-prequalified fourth-generation RDTs was performed using a bivariate random-effect model.

**Results:** 1,932 records were screened, of which 31 diagnostic accuracy studies from 19 countries were included. 15 studies used US-only approved products, 12 used WHO-prequalified products and four used commercially discontinued products. The pooled sensitivity of WHO-prequalified Ag/Ab RDTs for acute HIV infection (AHI) was 94% (95% CI: 86%-99%). An RNA threshold of ≥ 1,000,000 copies/mL was used as a proxy for high viraemia and used as a cut-off for the following analyses. The cut-off based analysis is considered more suited to decision-making, as it focuses on cases most likely to be associated with higher viraemia and greater potential for detection during the p24 Ag window. When using enzyme immunoassay (EIA) as the reference standard, the pooled p24 Ag sensitivity was 76% (95% CI: 62%-88%), and the pooled p24 Ag sensitivity when using nucleic acid amplification test (NAAT) as the reference standard was 75% (95% CI: 41%-97%). In the general population, the pooled sensitivity for p24 antigen detection was 77% (95% CI: 60%-92%). Amongst risk populations, only three studies had available raw data, and the pooled sensitivity was 62% (95% CI: 10%-97%). In plasma and serum specimens, pooled p24 Ag sensitivity was 74% (95% CI: 57%-88).

**Discussion:** Collectively, these findings indicate that WHO-prequalified fourth-generation Ag/Ab RDTs can function as a scalable frontline screening tool, particularly in low- and middle-income countries, while offering incremental holistic detection through p24 Ag. Their effective deployment, however, depends on maintaining standard algorithm safeguards, including repeat testing and targeted laboratory referral when acute infection is suspected.

**Conclusions:** Results from this meta-analysis support the use of WHO-prequalified fourth-generation Ag/Ab RDTs for general population screening. From a programmatic perspective, the added value of WHO-prequalified fourth-generation RDTs lies in their ability to combine rapid, decentralized access to testing, with incremental yet impactful improvements in holistic detection.

## 1. INTRODUCTION

HIV remains a major global health challenge: an estimated 40.8 million people were living with HIV and around 1.3 million people acquired HIV in 2024 (1). Despite substantial progress in expanding access to testing and treatment, important diagnostic gaps persist. Although around 87% of people living with HIV were aware of their status, an estimated 5.3 million people remained undiagnosed, and 630,000 people died from AIDS-related illnesses in 2024 (1). Diagnostics have become the backbone of HIV prevention, treatment and retention in care, and play a pivotal role in achieving the 95-95-95 UNAIDS targets (2). Accurate and timely diagnosis enables prompt initiation of antiretroviral therapy (ART), improving clinical outcomes and reducing morbidity (2,3). This is particularly critical in acute HIV infection, when viral loads are highest, and individuals contribute disproportionately to onward transmission (4,5), because the viruses during the acute phase of infection are known to be more infectious (6). By enabling earlier detection, antigen (Ag) testing can help close this diagnostic window, support faster linkage to ART and prevention services, and ultimately reduce onward transmission (4). In addition, knowledge of HIV serostatus facilitates effective prevention strategies, including pre-exposure prophylaxis (PrEP) for HIV-negative partners and treatment as prevention among key and at-risk populations (3).

To effectively reach populations who need it most, diagnostic technologies must be cost-effective, robust, user-friendly and suitable for settings with limited infrastructure. Amongst the different test techniques available, rapid diagnostic tests (RDTs) play a central role in scaling-up HIV testing (7). The introduction of RDTs provides important operational advantages, facilitating decentralized HIV testing and subsequently improving access to services for hard-to-reach populations (8–11). It also supports cost-effective HIV screening and targeted interventions such as the prevention of mother-to-child transmission and testing services for key populations (7,12). However, the deployment of these tests is contingent upon the assurance that they are of high-quality. To help ensure the quality of diagnostic tests, WHO launched the prequalification of *in vitro* diagnostics (IVD) in 2010 to support procurers’ source prequalified IVD with confidence that these products are quality-assured and suitable for their intended settings of use (13).

One limitation of earlier RDTs was their inability to detect acute HIV, with limited ability to p24 antigen, an early marker of HIV infection, which extended the diagnostic window to as long as two months. The introduction of fourth-generation RDTs, which incorporate p24 Ag detection, was intended to improve earlier diagnosis by shortening the detection window to approximately two-to-three weeks (14). Earlier detection through p24 Ag-enabled RDTs can also strengthen timely linkage to prevention and treatment services and support more comprehensive care pathways.

Against this backdrop, this review builds on a recent WHO systematic review that assessed the performance of fourth-generation HIV RDTs with discriminatory detection between p24 Ag and antibodies (Ab) (15). The meta-analysis evaluated class-level performance of fourth-generation RDTs, including products that had not received WHO-prequalification or that had been discontinued. To more accurately capture the current diagnostic landscape, this systematic review extends previous work by evaluating the performance of fourth generation RDTs at the individual product level, focusing exclusively on WHO prequalified fourth generation Ag/Ab assays.

## 2. METHODS

This systematic review was conducted according to Preferred Reporting Items for Systematic Reviews and Meta-Analyses (PRISMA) guidelines and the Cochrane Handbook for Systematic Reviews of Diagnostic Test Accuracy (DTA) reviews (16,17).

### 2.1. Eligibility criteria

Studies were selected according to predefined inclusion and exclusion criteria (**Table 1**).

**Table 1.** Overview of PICO criteria.

| PICO | Inclusion Criteria | Exclusion Criteria |
| --- | --- | --- |
| **Population** | Individuals aged $\geq 12$ years | Children $< 12$ years<br>Studies evaluating NAT to diagnose children as part of early infant diagnosis or monitoring HIV treatment |
| **Intervention** | Studies investigating the use of fourth-generation HIV 1/2 Ag/Ab RDTs which have received WHO-prequalification for the detection of acute/early HIV infection (AEHI) | Non-WHO-prequalified RDTs or NAT products (e.g., products which may have receive US FDA approval only)<br>Discontinued/non-commercially available products |
| **Comparator** | Studies using fourth-generation HIV 1/2 enzyme immunoassays (EIAs) with discriminatory detection of HIV Ab and p24 Ag<br>HIV-1 NAT can be used as reference standard (if EIA data not available) | Other tests |
| **Outcomes** | Diagnostic performance of fourth-generation HIV 1/2 Ag/Ab RDTs which have received WHO-prequalification | Outcomes not focusing on the diagnostic performance of fourth-generation HIV 1/2 Ag/Ab RDTs which have received WHO-prequalification |
| **Study design** | Randomized control trials, cohort studies, cross-sectional studies and case control studies | Reviews, case studies and non-clinical studies, reports from manufacturers, instructions for use and opinion papers |
| **Timeframe** | Studies published between 1 January 2010 and 31 December 2025 | Studies published before 1 January 2010<br>Studies published after 31 December 2025 |
| **Other** | Studies with available raw data | Studies where raw data for p24 Ag and RNA was not available |

### 2.2. Search strategy and selection

#### 2.2.1. Information sources and study selection process

Two reviewers (MN, RH) independently performed title and abstract screening across six online databases (6-12 January 2026): ClinicalTrials.gov, Cochrane Library, PubMed, Ovid MEDLINE, Scopus, and Web of Science. Database specific combinations of key words and MeSH terms were applied (e.g., “acute HIV,” “antigen-antibody,” “rapid diagnostic tests”), and searches were restricted to a date range from 1 January 2010 to 31 December 2025. Language restrictions were not applied, with non-English studies translated on DeepL. Full search strategies are provided in Appendix S1. Identification of additional records was conducted via manual reference, citation searching and conference abstract searching.

All records were imported into Rayyan, a web-based tool designed to streamline and accelerate the screening and study selection process, where duplicates were removed prior to screening. Record counts and exclusion reasons at each stage were also documented using Rayyan. Remaining full-text articles were assessed for eligibility by the same two independent reviewers, documenting reasons for full-text exclusion.

#### 2.2.2. Data extraction

Data extraction was performed by two independent reviewers (MN, RH) using a pre-determined Excel template and assessed for final outcomes. Discrepancies were resolved by consensus. The extracted data included raw diagnostic accuracy metrics for HIV p24 Ag and HIV Ab (true positive, false positive, false negative, true negative, and study meta-data including but not limited to study identification, first author, publication year, country, population, setting, study design, sample size, specimen type, relevant cut-offs, name and type of index and reference tests used. Studies without available raw data were excluded.

### 2.3. Risk of bias assessment

The risk of bias in diagnostic accuracy studies was assessed across four domains (patient selection, index test, reference standard, and flow and timing), using the QUADAS-2 tool (18).

### 2.4. Data synthesis and analysis

#### 2.4.1. Diagnostic accuracy

The primary study outcome was the diagnostic performance of fourth-generation HIV 1/2 Ag/Ab RDTs which have received WHO-prequalification (Appendix S2), as compared to other molecular or serological lab-based assays. Acute HIV infection (AHI) was defined as a positive p24 Ag or HIV RNA test in the presence of seronegative or discordant antibody results. Studies that did not report sufficient raw data to calculate sensitivity were excluded from the analysis.

The following pooled estimates were computed as part of the meta-analysis: sensitivity, specificity, positive predictive value (PPV), negative predictive value (NPV), and proportion of false positive and false negative cases with their 95% confidence intervals.

A Bayesian random-effects model using specific parametrization for diagnostic meta-analysis was prepared according to literature best practice. The model was built in reference statistics analysis language R (v. 4.5.2), using validated packages recognized as accurate specifically in the context of Bayesian diagnostic meta-analyses. The model performs bivariate analysis using nested Laplace approximations, with half-Cauchy priors (19,20). Outputs are shown as forest plots, to allow for visual inspection of study heterogeneity. The analysis was run twice: once including all RNA values and once applying a cut-off of RNA ≥ 1,000,000 copies/mL. The forest plots presented here reflect the ≥ 1,000,000 copies/mL analysis only.

Studies lacking quantitative viral load data contributed to overall Ag/Ab sensitivity estimates but could not provide sufficient data for p24 Ag analyses and were therefore excluded. Pooled estimates were completed based on comparable reference standards or HIV testing strategies and algorithms.

#### 2.4.2. Rationale for viral load–based analytical cut-offs

Given the known discordance between HIV RNA detectability and p24 Ag expression during early infection, the sensitivity analyses were run twice: (i) without a viral load cut-off, and (ii) applying an RNA threshold of ≥ 1,000,000 copies/mL as a proxy for high viraemia (21–24). The cut-off based analysis is considered more suited to decision-making, as it focuses on cases most likely to be associated with higher viraemia and greater potential for detection during the p24 Ag window (25,26).

This cut-off was selected for four reasons: (i) biological plausibility, as peak viraemia during acute infection commonly exceeds this level and coincides with maximal p24 Ag expression; (ii) reduction of between-study heterogeneity by restricting analyses to a more homogeneous biological subgroup; (iii) improved interpretability, as p24 Ag detection is expected under high-viraemia conditions, avoiding conflation of biological limitations with analytical or operational shortcomings; and (iv) analytical intent, as the cut-off was applied only as a stratification variable rather than as a definition of HIV positivity or an exclusion criterion. All eligible studies were retained in the review, and high-viraemia analyses were conducted only when viral load data were available.

## 3. RESULTS

2,472 records were identified through the initial search, with 1,932 unique records remaining after deduplication. Following title and abstract screening, 109 full text articles were assessed for eligibility, of which 31 studies focused on diagnostic accuracy were included in the qualitative synthesis (**Figure 1**). 12 studies included WHO-prequalified fourth-generation RDTs, of which only 10 provided sufficient raw data to support quantitative synthesis (27–36). One study was included only for the overall Ag/Ab performance, but not for p24 Ag analysis due to the absence of RNA cut-off (27).

**Figure 1.**
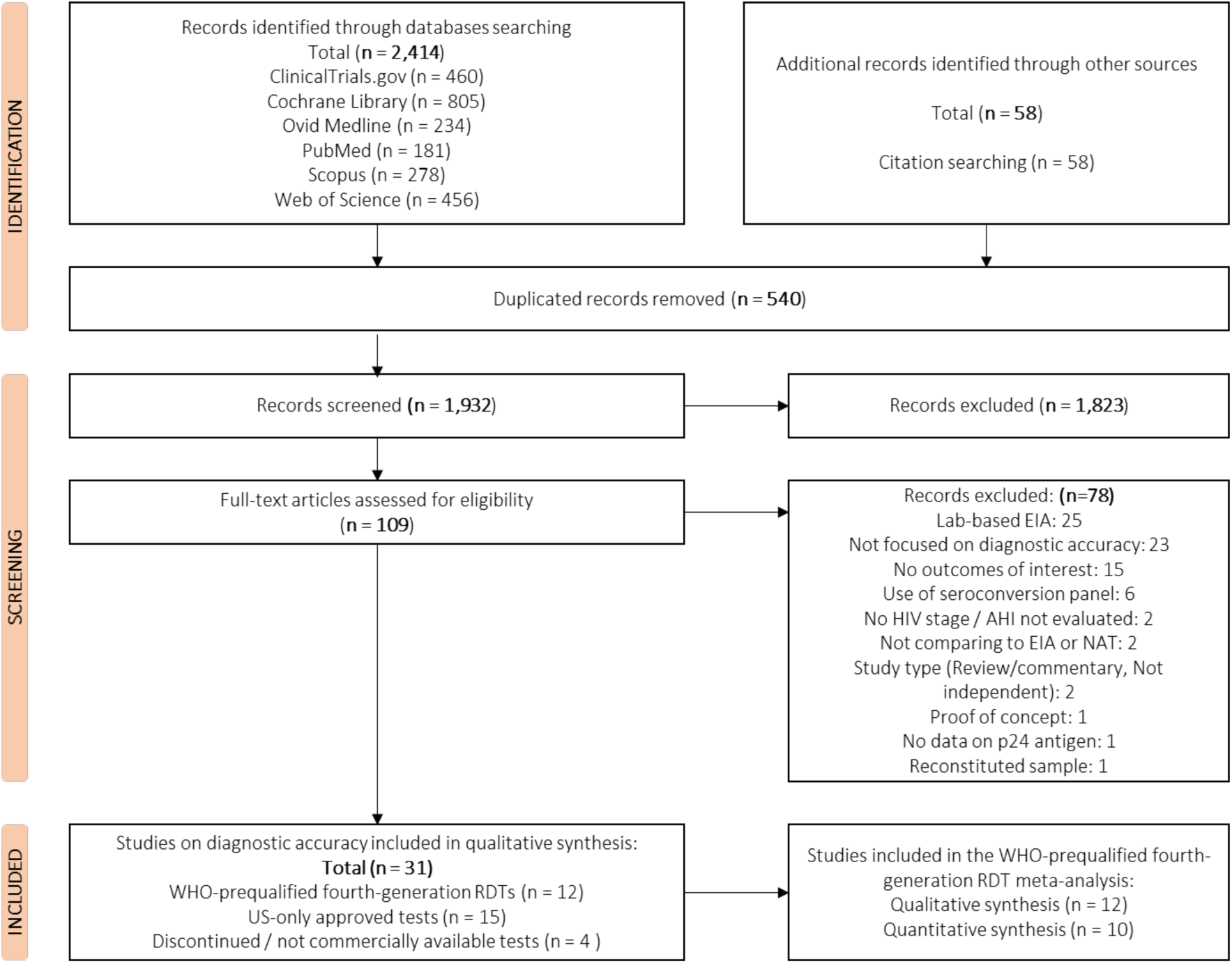
PRISMA flowchart of included and excluded records.

### 3.1. Study characteristics

Diagnostic accuracy was reported in 31 studies, the characteristics of which are summarized in **Table 2**. The populations studied included adult individuals at high-risk of HIV infection (45% of studies), individuals with existing diagnoses (13%), general population (10%), PrEP users (10%), and hospital patients (10%). The final 12% of studies investigated archived specimens from laboratory-based case-control studies. Studies were conducted in 19 countries, mainly in lab settings (55%) and clinics (26%). The primary specimen types used were plasma (40%) and serum (35%), while fingerstick, venous blood and whole blood were only used in a small share of studies (13%, 8% and 3%, respectively).

**Table 2.**
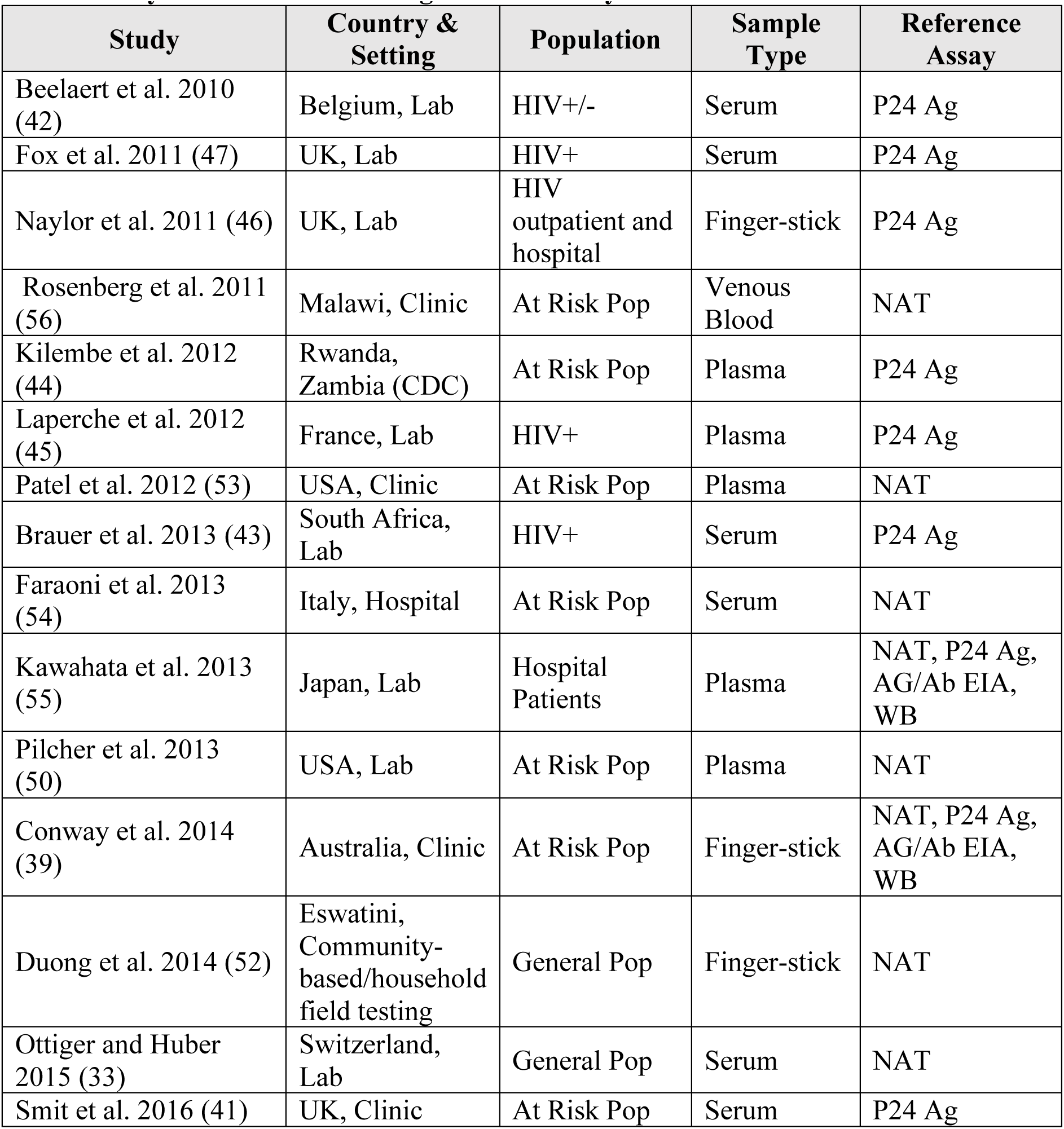
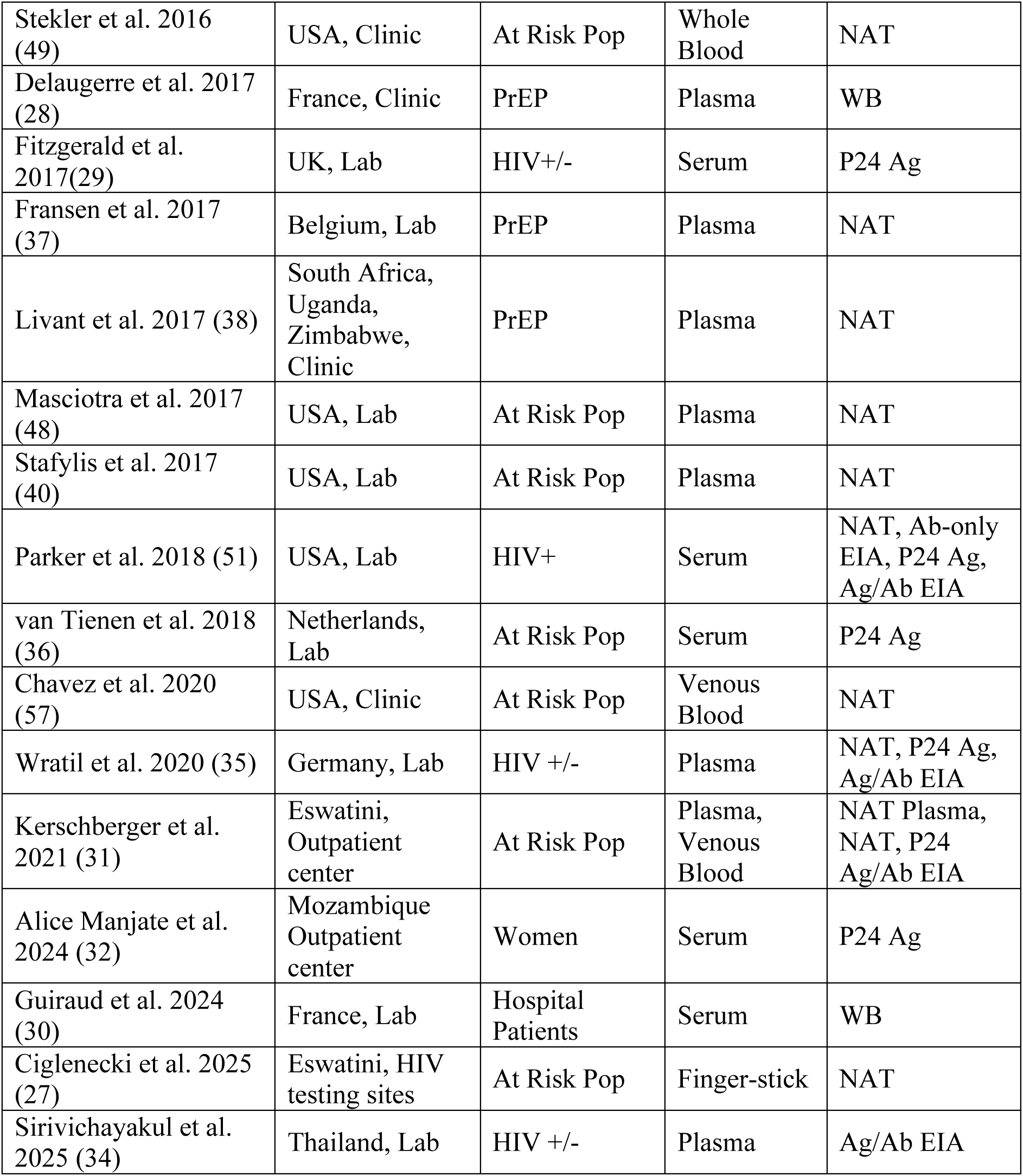
Key characteristics of diagnostic accuracy studies.

Of the included studies, 12 used WHO-prequalified fourth-generation RDTs (27–38), 15 used US-approved only products (39–53), and four used commercially discontinued products (54–57). The studies evaluating non-WHO-prequalified fourth-generation HIV RDTs as index tests were excluded, as such assays are not eligible for WHO-endorsed testing algorithms or large-scale public health procurement processes in low- and middle-income countries (LMICs). Limiting inclusion to WHO-prequalified tests ensured that pooled estimates reflect performance under conditions relevant to current policy, implementation, and regulatory standards.

### 3.2. Risk of bias assessment

QUADAS-2 was used to appraise risk of bias for diagnostic accuracy studies (Appendix S3). Across all studies, risk of bias was found to be low for reference standard and flow and timing domains. For patient selection, risk of bias was frequently found to be high (in 70% of studies), largely due to case-control study designs and non-consecutive sampling. Risk of bias for the index test was low in 36%, high in 16% and unclear in 48%, primarily due to insufficient reporting on whether test administrators were blinded to reference standard results.

### 3.3. Meta-analysis on WHO-prequalified fourth-generation Ag/Ab RDT

Meta-analysis of 10 studies using WHO-prequalified fourth-generation Ag/Ab RDTs had a pooled sensitivity of 94% (95% CI: 86%-99%), with moderate heterogeneity across studies. The majority of studies demonstrated strong performance, with eight out of ten studies clustering around a moderate-to-high sensitivity (84% to 99%), while two out of ten studies presented lower estimates (Appendix S4). All subsequent p24 Ag sensitivity analyses apply an RNA cut-off ≥ 1,000,000 copies/mL, and the findings should be interpreted in that context.

The pooled p24 Ag sensitivity was at 73% (95% CI: 55%-88%), indicating a moderate ability to correctly identify HIV-positive individuals (**Figure 2**). Five out of nine studies clustered around a moderate sensitivity (65% to 72%), with three studies reporting higher sensitivity estimates (85% to 87%). One study was a clear low outlier and contributed disproportionately to between-study heterogeneity (31). Studies with wide confidence intervals, most notably Manjate et al., (2024) (32), were characterized by very small sample sizes, indicating imprecision rather than systematically reduced test performance.

**Figure 2.**
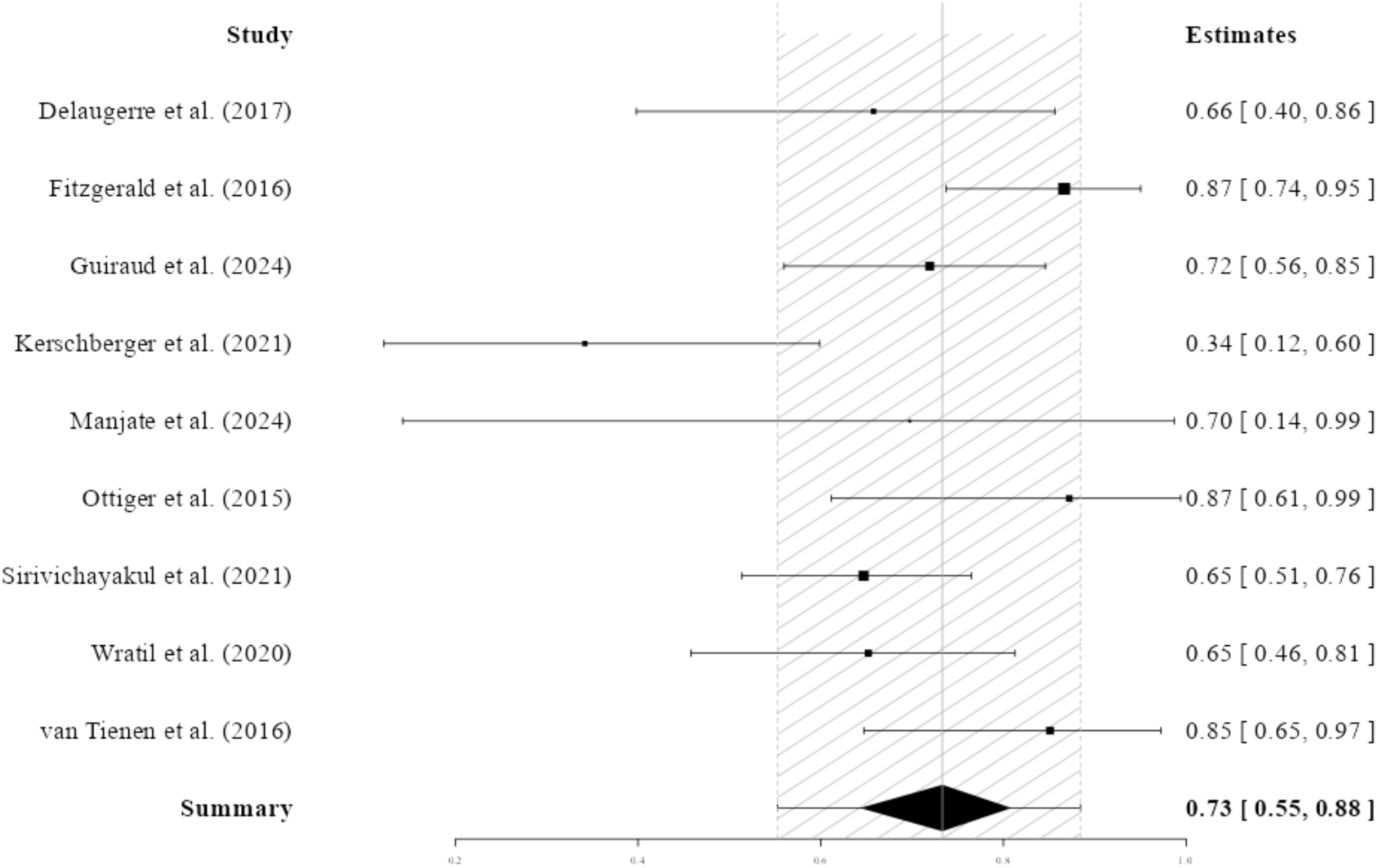
Overall diagnostic sensitivity for p24 Ag detection of WHO-prequalified HIV Ag/Ab fourth-generation RDTs.

When using EIAs as the reference standard, the pooled p24 Ag sensitivity was 76% (95% CI: 62%-88%) (**Figure 3**). Sensitivity estimates were consistent across studies, with five out of eight studies clustering around a moderate-to-high range, with a sensitivity of 66% to 84%.

**Figure 3.**
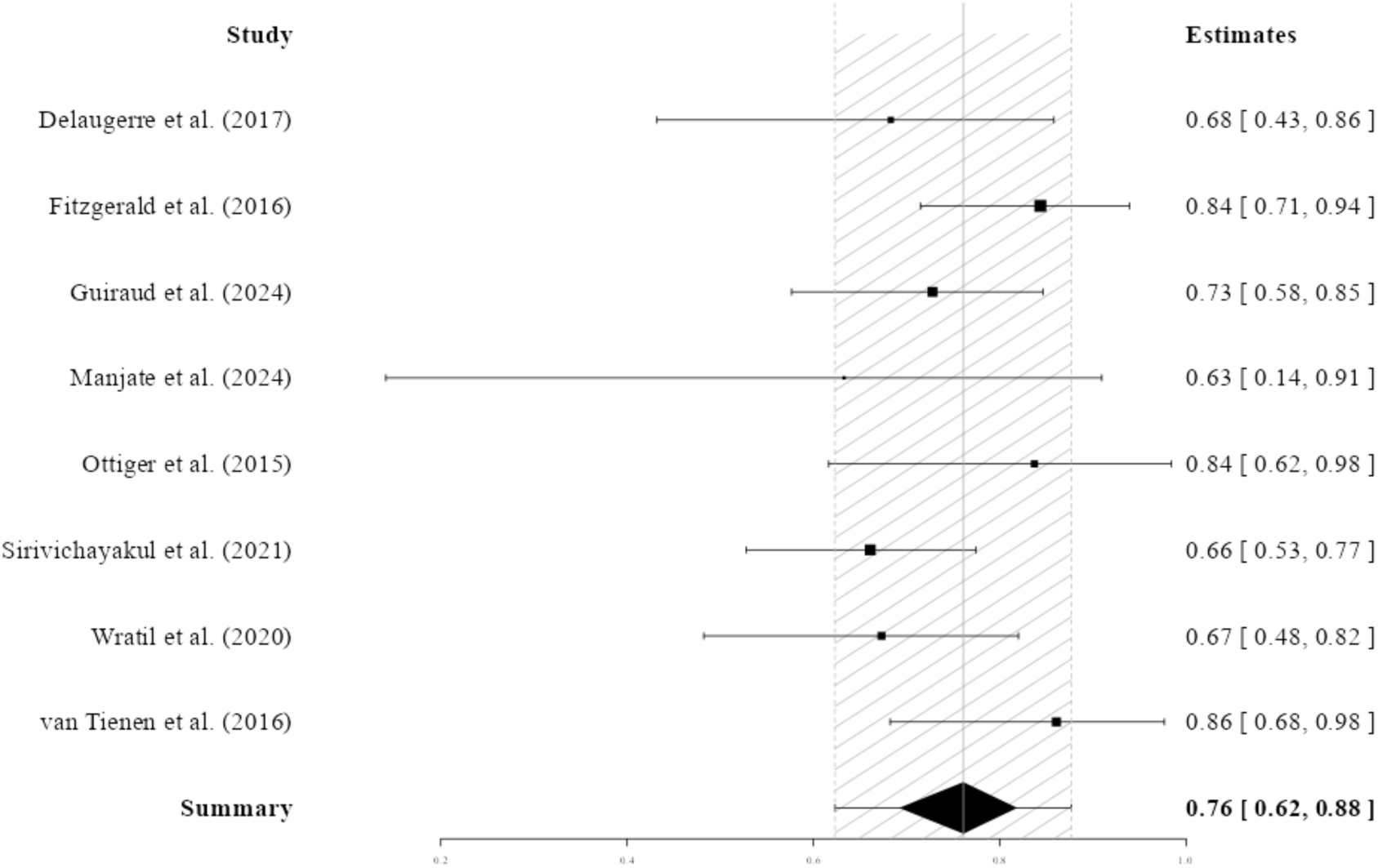
Diagnostic sensitivity for p24 Ag detection of WHO-prequalified HIV Ag/Ab fourth-generation RDTs compared to EIA.

Residual heterogeneity was largely attributable to differences in study size and design rather than divergent assay performance, with wider confidence intervals again observed in studies with smaller sample sizes.

When restricting analyses using a cut-off of RNA ≥ 1,000,000 copies/mL and excluding studies without quantitative data available for viral load (27,37,38), the pooled p24 Ag sensitivity compared to NAAT was 75% (95% CI: 41%-97%) (**Figure 4**).

**Figure 4.**
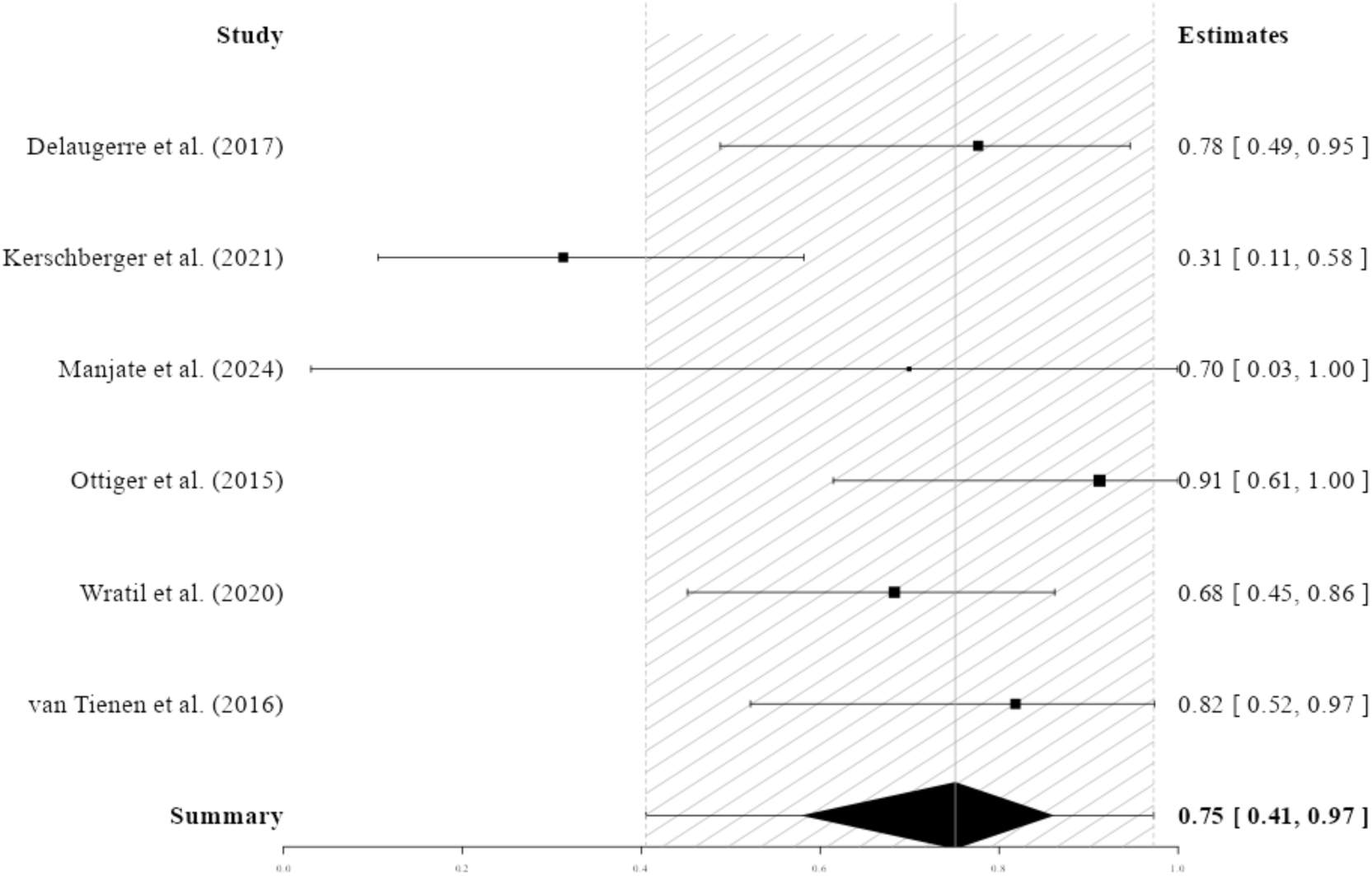
Diagnostic sensitivity of p24 antigen detection compared to NAAT (cut-off at RNA ≥ 1,000,000 copies/mL).

Sensitivity estimates ranged from 31% to 91% across six studies. Three studies reported high sensitivity (78% to 91%), while one study remained a clear low outlier (31). The wide confidence intervals observed in this analysis likely reflect both reduced sample sizes following viral load stratification and residual biological heterogeneity within high-viraemia populations (32).

In the general population, the pooled sensitivity for p24 Ag detection was 77% (95% CI: 60%-92%) (**Figure 5**). Sensitivity estimates across studies ranged from 66% to 85%, with the majority of studies (five out of six) clustering between 70% and 84%, indicating consistent performance across general population settings. Limited variability was observed, largely driven by a single small-sample study (32), with otherwise stable estimates across studies.

**Figure 5.**
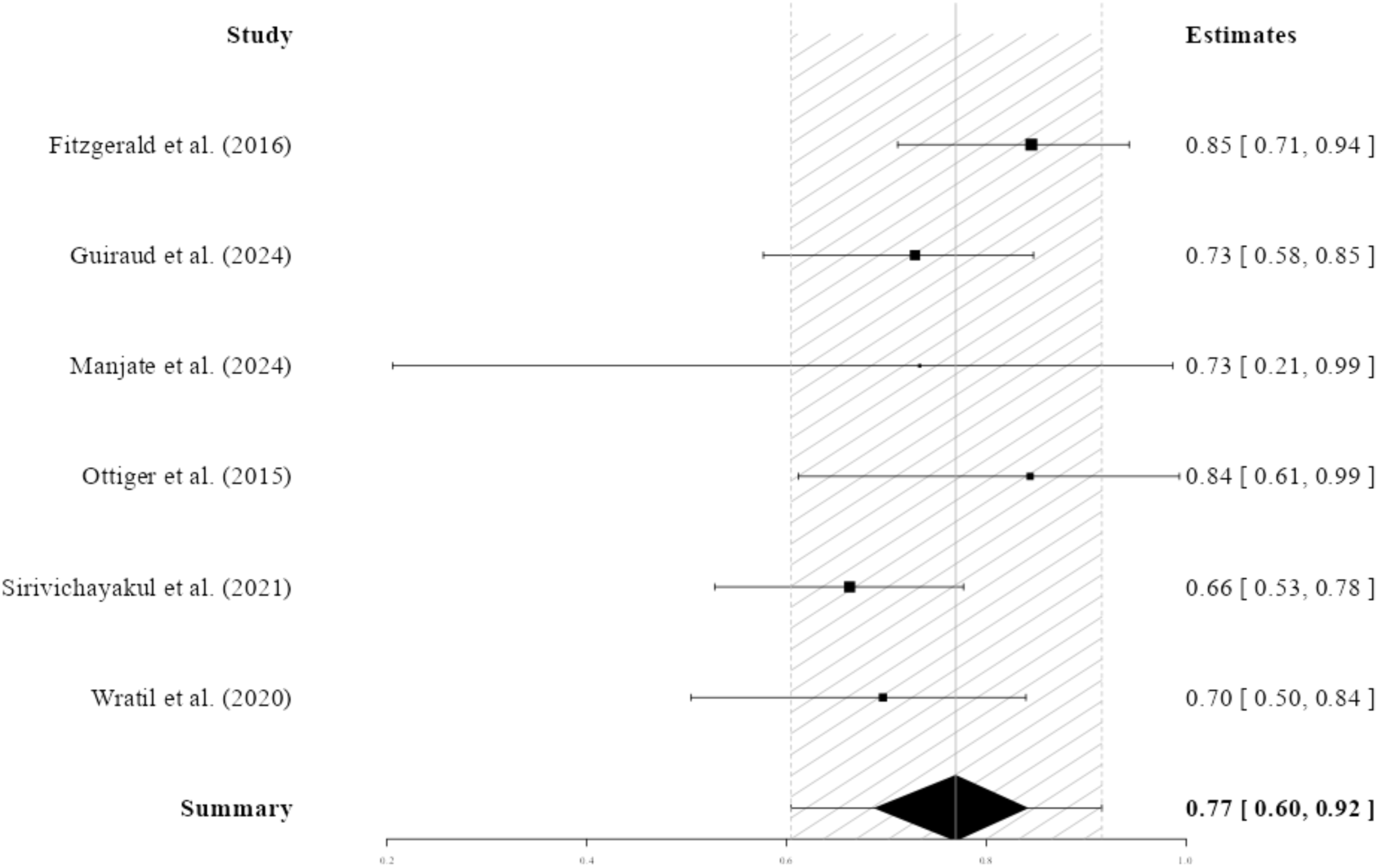
Diagnostic sensitivity for p24 Ag detection of WHO-prequalified HIV Ag/Ab fourth-generation RDTs in the general population.

Amongst risk populations, only three studies had available raw data. Amongst these, the pooled sensitivity was 62% (95% CI: 10%-97%), but the estimate is highly imprecise as reflected by the wide confidence interval (Appendix S5). Furthermore, some individuals within the included study populations had been receiving PrEP or ART, which can influence the immunological responses and may affect p24 Ag levels and test sensitivity. As such, this data should be used as directional evidence and should not be used to compare performance between population types.

In plasma and serum specimens, pooled p24 Ag sensitivity was 74% (95% CI: 57%-88). Eight out of nine studies clustered in a mid-to-high performance range (65% to 87%). One low-performing study and wider confidence intervals in studies with small sample sizes contributed to heterogeneity (31,32), suggesting context-specific effects and imprecision rather than systematic assay limitations (Appendix S6).

Across all analyses, heterogeneity in p24 Ag sensitivity estimates was primarily driven by infection stage, sample size, and study design rather divergent test performance. **Table 3** provides a consolidated summary of pooled sensitivity estimates across reference standards, viral load strata, and population and specimen subgroups.

**Table 3.**
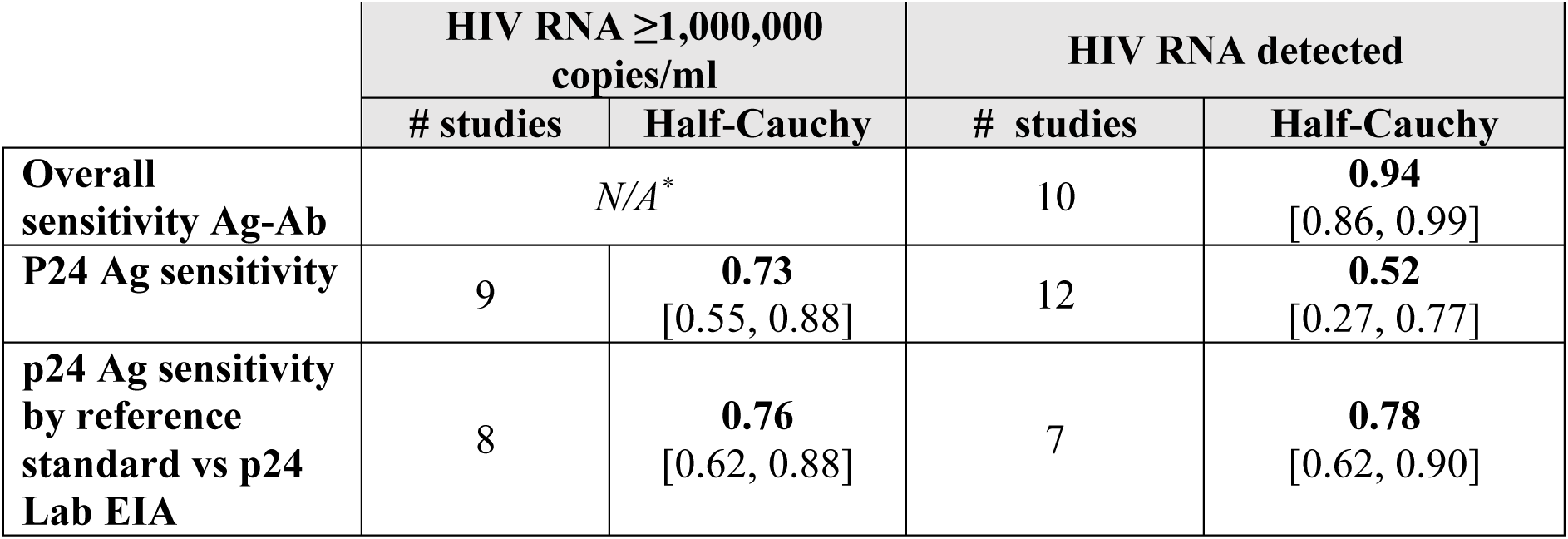
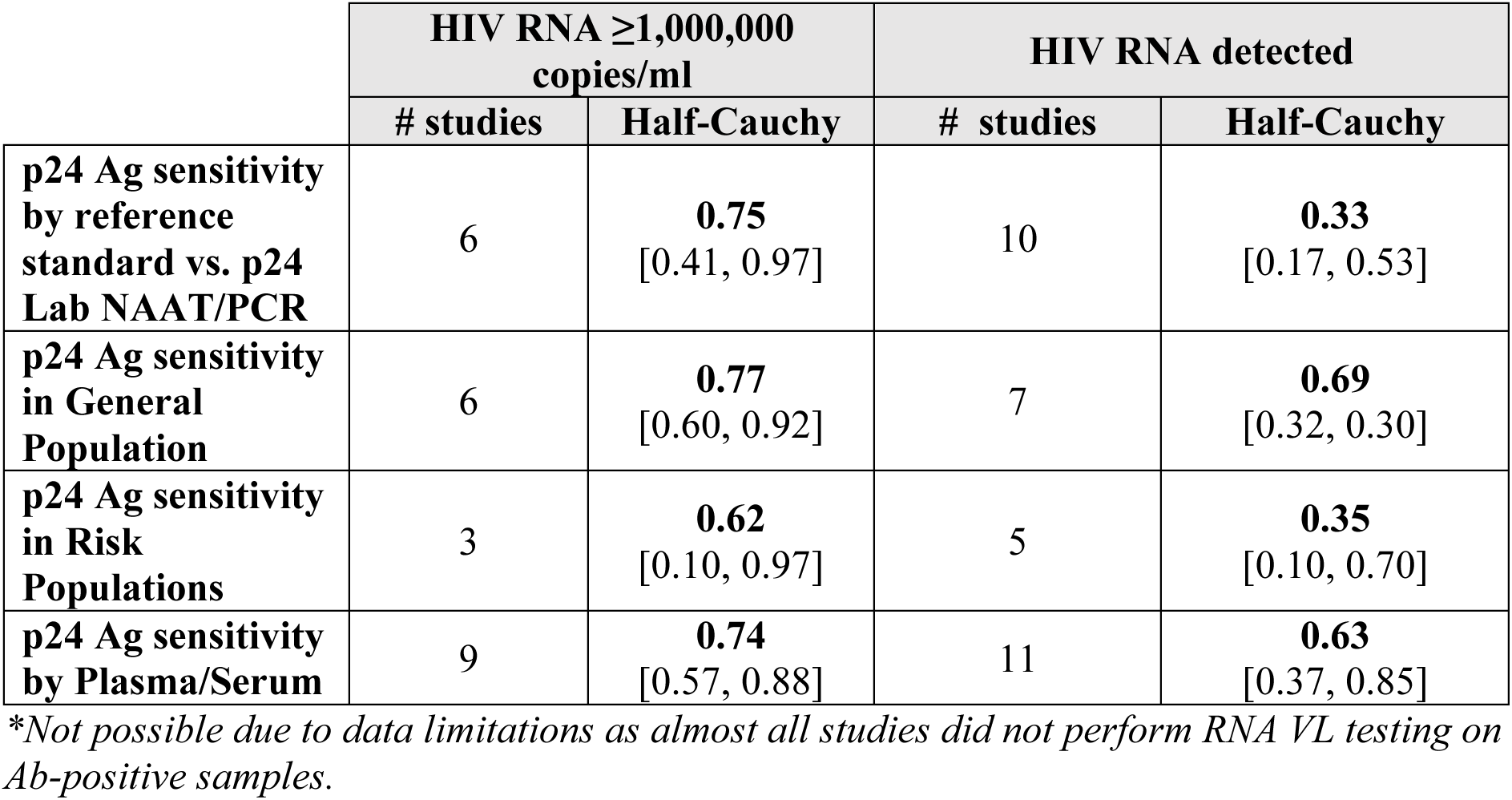
Summary of pooled diagnostic accuracy estimates for WHO-prequalified fourth-generation HIV Ag/Ab RDTs.

| | HIV RNA $\geq 1,000,000$ copies/ml | | HIV RNA detected | |
| --- | --- | --- | --- | --- |
|  | # studies | Half-Cauchy | # studies | Half-Cauchy |
| **Overall sensitivity Ag-Ab** | *N/A\** |  | 10 | **0.94**<br>[0.86, 0.99] |
| **P24 Ag sensitivity** | 9 | **0.73**<br>[0.55, 0.88] | 12 | **0.52**<br>[0.27, 0.77] |
| **p24 Ag sensitivity by reference standard vs p24 Lab EIA** | 8 | **0.76**<br>[0.62, 0.88] | 7 | **0.78**<br>[0.62, 0.90] |
|  | # studies | Half-Cauchy | # studies | Half-Cauchy |
| **p24 Ag sensitivity by reference standard vs. p24 Lab NAAT/PCR** | 6 | **0.75**<br>[0.41, 0.97] | 10 | **0.33**<br>[0.17, 0.53] |
| **p24 Ag sensitivity in General Population** | 6 | **0.77**<br>[0.60, 0.92] | 7 | **0.69**<br>[0.32, 0.90] |
| **p24 Ag sensitivity in Risk Populations** | 3 | **0.62**<br>[0.10, 0.97] | 5 | **0.35**<br>[0.10, 0.70] |
| **p24 Ag sensitivity by Plasma/Serum** | 9 | **0.74**<br>[0.57, 0.88] | 11 | **0.63**<br>[0.37, 0.85] |

*\*Not possible due to data limitations as almost all studies did not perform RNA VL testing on Ab-positive samples.*

## 4. DISCUSSION

The findings from this review indicate high overall Ag/Ab sensitivity (94%, 95% CI: 86%-99%), with most studies demonstrating consistently strong performance. For early infection yield, pooled p24 Ag sensitivity was moderate to high (73%, 95% CI: 55%-88%), with generally consistent estimates across studies but influenced by a small number of low-precision studies (e.g., small sample sizes or single outliers). When stratified by reference standard, p24 Ag sensitivity was more stable against EIA (76%, 95% CI: 62%-88%) and slightly more variable against NAAT (75%, 95% CI: 41%-97%), reflecting the earlier detectability of RNA relative to antigen and the biologically earlier benchmark set by NAAT.

Subgroup analyses suggested predictable performance in general-population screening (77%, 95% CI: 60%-92%), whereas estimates in risk populations were highly imprecise due to limited available data (62%, 95% CI: 10%-97%). p24 Ag sensitivity in plasma/serum was 74% (95% CI: 57%-88%), indicating that specimen type may influence early detection yield.

Collectively, these findings indicate that WHO-prequalified fourth-generation Ag/Ab RDTs can function as a scalable frontline screening tool, offering incremental holistic detection through p24 Ag provided programs maintain standard algorithmic safeguards, such as repeat testing and targeted laboratory referral when acute infection is suspected. Although p24 Ag detection does not identify all acute infections, the moderate pooled sensitivity demonstrates that WHO-prequalified fourth-generation Ag/Ab RDTs can meaningfully shorten the diagnostic window compared to an antibody-only approach, enabling earlier diagnosis and faster linkage to care (5). This added early detection of acute HIV is particularly valuable in decentralized testing models (58), where rapid point-of-care results support immediate counselling, same-day referral and timely treatment initiation. In these settings, expanding access beyond laboratory-dependent pathways can improve coverage among underserved populations and help reduce inequities in diagnosis and health outcomes (59).

The findings are also highly relevant from a health economic perspective. HIV testing strategies that expand access at scale and support rapid diagnosis and prompt linkage to ART have consistently been shown to be cost-effective at the population level. The economic and public health value derives from enabling earlier ART initiation, reducing onward transmission through earlier viral suppression, and lowering longer-term treatment and care costs that arise from delayed or laboratory-dependent testing pathways (60–66). Within this context, the combined advantages of scalability, decentralized delivery and more timely detection reinforce the role of WHO-prequalified fourth-generation Ag/Ab RDTs as a practical tool for strengthening HIV testing strategies, while preserving essential algorithm safeguards such as repeat testing and targeted laboratory referral when acute infection is suspected.

A key contribution of this review is that it provides decision-relevant estimates anchored specifically in WHO-prequalified products, the assays most likely to be prioritized for national procurement and implementation. By restricting the analysis to WHO-prequalified fourth-generation RDTs and synthesising evidence at the product-level, the review avoids conflating performance expectations with assays that are ineligible for many procurement pathways or that do not meet comparable quality standards. The WHO-prequalification program evaluates diagnostic products for performance, manufacturing quality and suitability for use in LMICs, ensuring they meet rigorous, globally recognized benchmarks (67).

Although originally established to support United Nations procurement agencies, WHO-prequalification listings are now widely used by national programs and procurement bodies worldwide to guide vendor selection and secure quality-assured products (67–69). Focusing the analysis on WHO-prequalified fourth-generation Ag/Ab RDTs was intended to enhance the practical relevance of the findings for procurement and policy-decision making, as these assays are commonly incorporated into national testing algorithms.

This review has several strengths that can help inform HIV testing strategies. First, the evidence is synthesized at the product-level and restricted to WHO-prequalified fourth-generation Ag/Ab RDTs. This approach ensures that the pooled estimates reflect assays that meet a consistent quality benchmark and are the products most likely to be procured and implemented within national programs. Second, a random-effects, Bayesian DTA meta-analytic approach was used to account for heterogeneity, and the study examined key sources of variability through multiple stratifications, helping to contextualize the pooled performance estimates and support a more nuanced programmatic interpretation. Finally, by explicitly distinguishing overall Ag/Ab performance from p24-specific Ag performance, the review offers clearer insight into the incremental value contributed by antigen detection.

Despite these strengths, the present study has several limitations. First, not all eligible studies could be included in the quantitative analysis because extractable raw data were limited, particularly for p24 Ag-specific calculations. Even among studies with available raw data, variability in reporting clarity and methodological detail may have reduced the precision of data interpretation. Furthermore, definitions of AHI were applied inconsistently across studies, limiting comparability and complicating the assessment of diagnostic accuracy. Most included articles did not classify AHI according to the classical Fiebig staging system(70).

For the purposes of this review, AHI was defined by the detection of HIV p24 antigen, which may be detectable from the late Fiebig I stage through Fiebig III. Some analyses included only a small number of studies, most notably the stratified analysis by risk population, for which only three studies met inclusion criteria. Consequently, the resulting uncertainty intervals were wide, limiting the generalizability of these pooled estimates. In addition, most included studies were conducted under laboratory or otherwise controlled conditions, which may overestimate performance compared to routine field settings, where operator variability and environmental factors can affect results. The analysis did not adjust for operational differences across individual studies (e.g., specimen handling, testing conditions).The type of specimen also represents an additional source of selection bias. Differences between venous blood, plasma, serum, and fingerstick whole blood are well documented, and comparisons across specimen types may yield inconsistent results.

Finally, comparisons between NAT and fourth-generation Ag/Ab RDTs reflect differences in detection thresholds. NAT can detect infection at very low viral loads and p24 antigen levels that are often below the detection limits of fourth-generation Ag/Ab RDTs. Such biological variability inevitably affects systematic reviews and meta-analyses, as heterogeneity in infection stage and marker concentrations cannot be precisely determined or controlled.

Consequently, sensitivity estimates may underestimate the true performance of fourth-generation Ag/Ab RDTs for detecting acute HIV infection where comparisons are often made against reference methods with lower detection thresholds.

Future research should prioritize strengthening the evidence base in key populations and other higher-risk groups, where data remains limited. The robustness of future reviews would also benefit from more consistent reporting of clearly extractable p24 Ag raw data, including specified cut-offs. This would enable more reliable product-level analyses. Finally, additional research is needed in LMICs, where these testing use cases are often most operationally and clinically relevant. This will be critical for ensuring that performance expectations and implementation recommendations are applicable to the settings in which the expansion of testing programs is most needed.

From a pragmatic program perspective, these results support the inclusion of WHO-prequalified fourth-generation Ag/Ab RDTs as a reliable first-line screening tool within national testing algorithms, with the additional advantage of enhanced early infection detection. In decentralized testing environments, the incremental p24 Ag yield can meaningfully enhance decision-making in situations requiring rapid action and minimal laboratory support. Where NAAT is unavailable or unaffordable, fourth-generation Ag/Ab RDTs provide a practical means of partially narrowing the acute-infection diagnostic gap, provided that testing algorithms maintain safeguards (e.g., repeat testing) to rule out very early infection. Indeed, the integration of fourth-generation HIV Ag/Ab rapid diagnostic tests into routine testing algorithms could meaningfully strengthen early HIV detection in resource-constrained settings where transmission during acute infection continues to undermine epidemic control. While operational realities, including cost, supply chain stability, workforce training and algorithm complexity, must be carefully managed, the findings from this review suggest that strategic, targeted deployment offers the most pragmatic pathway forward. Aligning implementation with national HIV Testing Services frameworks, high-yield entry points (e.g., key populations, STI clinics, index testing, and high-incidence settings), and robust quality assurance systems can maximize public health impact while preserving program efficiency. Importantly, earlier diagnosis through improved detection of acute HIV infection has implications beyond individual clinical outcomes, enabling faster treatment initiation and reducing onward transmission at the population level. Future implementation research and cost-effectiveness evaluations will be critical to inform national scale-up decisions, but the present findings underscore that optimizing early diagnosis represents a strategic opportunity to accelerate progress in high-burden, resource-limited settings.

## 5. CONCLUSIONS

Early detection of acute HIV infections provides substantial benefits at both the individual and population-levels. Early diagnosis enables prompt treatment initiation, which slows disease progression, reduce HIV-related morbidity and mortality, and lowers long-term healthcare costs by preventing advanced disease. For this, fourth-generation RDTs with discriminatory Ag/Ab detection play a central role in enabling this.

Findings from this meta-analysis support the use of WHO-prequalified fourth-generation Ag/Ab RDTs, provided that algorithmic safeguards are maintained, and national testing guidelines are followed. From a program perspective, their impact stems from the ability to pair rapid, decentralized testing with incremental gains in early infection detection. Even when only a subset of acute infections is identified, earlier diagnosis can expedite linkage to ART and facilitate timely prevention interventions, improving individual outcomes and advancing broader public health goals.

## Supporting information

All sumplemental tables

## Data Availability

Introduction: Diagnostics have become the fundamental backbone of HIV prevention, treatment and long-term retention in care, and are critical to achieving the 95-95-95 UNAIDS targets. To effectively reach underserved and remote populations, diagnostic technologies must be cost-effective, robust, user-friendly and suitable for settings with limited infrastructure. Among available testing modalities, rapid diagnostic tests (RDTs) play a central role in expanding HIV testing coverage. Earlier generations of RDTs were limited by their inability to detect acute HIV, with limited ability to detect p24 antigen (Ag), an early marker of HIV infection, which is expected to shorten the diagnostic window to two-to-three weeks. The introduction of fourth-generation RDTs, which detects both chronic and acute HIV infection through p24 Ag detection, was designed to ensure that the traditional diagnostic window of two-to-three months is shortened to approximately two-to-three weeks. However, integrating these assays into existing testing algorithms requires clear evidence that they meet high standards of quality and performance. This systematic review aims to assess the performance of WHO-prequalified fourth-generation Ag/Ab RDTs.
Methods: We performed a systematic search across six databases to identify studies evaluating Ag/Ab RDTs against laboratory reference standards in individuals aged 12 years and older, spanning 1 January 2010 to 31 December 2025. Outcomes were limited to measures of diagnostic accuracy. A meta-analysis focusing exclusively on WHO-prequalified fourth-generation RDTs was performed using a bivariate random-effect model.
Results: 1,932 records were screened, of which 31 diagnostic accuracy studies from 19 countries were included. 15 studies used US-only approved products, 12 used WHO-prequalified products and four used commercially discontinued products. The pooled sensitivity of WHO-prequalified Ag/Ab RDTs for acute HIV infection (AHI) was 94% (95% CI: 86%-99%). An RNA threshold of ≥ 1,000,000 copies/mL was used as a proxy for high viraemia and used as a cut-off for the following analyses. The cut-off based analysis is considered more suited to decision-making, as it focuses on cases most likely to be associated with higher viraemia and greater potential for detection during the p24 Ag window. When using enzyme immunoassay (EIA) as the reference standard, the pooled p24 Ag sensitivity was 76% (95% CI: 62%-88%), and the pooled p24 Ag sensitivity when using nucleic acid amplification test (NAAT) as the reference standard was 75% (95% CI: 41%-97%). In the general population, the pooled sensitivity for p24 antigen detection was 77% (95% CI: 60%-92%). Amongst risk populations, only three studies had available raw data, and the pooled sensitivity was 62% (95% CI: 10%-97%). In plasma and serum specimens, pooled p24 Ag sensitivity was 74% (95% CI: 57%-88).
Discussion: Collectively, these findings indicate that WHO-prequalified fourth-generation Ag/Ab RDTs can function as a scalable frontline screening tool, particularly in low- and middle-income countries, while offering incremental holistic detection through p24 Ag. Their effective deployment, however, depends on maintaining standard algorithm safeguards, including repeat testing and targeted laboratory referral when acute infection is suspected.
Conclusions: Results from this meta-analysis support the use of WHO-prequalified fourth-generation Ag/Ab RDTs for general population screening. From a programmatic perspective, the added value of WHO-prequalified fourth-generation RDTs lies in their ability to combine rapid, decentralized access to testing, with incremental yet impactful improvements in holistic detection.

## AUTHORS’ CONTRIBUTIONS

Conceptualization: MN, RH; Methodology: MN, RH; Formal analysis: MN, RH; Data curation: MN, RH, AL; Investigation: MN, RH; Writing – original draft: MN, RH; Writing – review & editing: MN, RH, VG, AP, AO, SS, AS, GR, MO; Visualization: AL; Supervision: MN; Project administration: MN; Funding acquisition: MN

## ACKNOWLEDGEMENTS

We sincerely thank Alexander Lyon for running the bivariate random-effect models in R using the Bayesian approach for diagnostic accuracy meta-analysis, and for his invaluable statistical guidance throughout the study.

## DISCLAIMER

Funders had no role in study design, analysis, or interpretation.

