## Supplementary material for "Should WHO-prequalified antigen-antibody fourth-generation rapid diagnostic tests be used to detect acute HIV infection? A systematic review and meta-analysis of diagnostic performance": All sumplemental tables

#### SUPPORTING INFORMATION

Additional information may be found under the Supporting Information tab for this article.

**Appendix S1. Full search strategies.**

| Database | Search Date | Search String | Date Limits Applied |
|---|---|---|---|
| PubMed | 12th January 2026 | ("HIV"[MeSH Terms] AND ("diagnosis"[MeSH Subheading] OR "diagnosis"[All Fields] OR "screening"[All Fields] OR "mass screening"[MeSH Terms] OR ("mass"[All Fields] AND "screening"[All Fields]) OR "mass screening"[All Fields] OR "early detection of cancer"[MeSH Terms] OR ("early"[All Fields] AND "detection"[All Fields] AND "cancer"[All Fields]) OR "early detection of cancer"[All Fields] OR "screen"[All Fields] OR "screenings"[All Fields] OR "screened"[All Fields] OR "screens"[All Fields] OR "diagnosis"[MeSH Terms]) AND ("rapid diagnostic test"[Title/Abstract] OR "RDT"[Title/Abstract] OR "point-of-care"[Title/Abstract] OR "POC"[Title/Abstract] OR "test"[Title/Abstract] OR "algorithm"[Title/Abstract]) AND ("fourthgeneration"[Title/Abstract] OR "4th generation"[Title/Abstract] OR "ag ab"[Title/Abstract] OR "antigen-antibody"[Title/Abstract] OR "Determine"[Title/Abstract] OR "Alere"[Title/Abstract]) AND ("acute hiv infection"[Title/Abstract] OR "AHI"[Title/Abstract] OR "early"[Title/Abstract] OR "recent"[Title/Abstract] OR "AEHI"[Title/Abstract])) AND (2010:2025[pdat]) | Publication year: 2010 - 2025 |
| Ovid Medline | 12th January 2026 | (HIV OR "acute HIV" OR "early HIV") AND (diagnosis OR screening) AND (sensitivity OR "diagnostic accuracy" OR "diagnostic performance" OR detection) AND ("fourth generation" OR "4th generation" OR Ag/Ab OR antigen- antibody) AND (test OR assay OR "rapid diagnostic test" OR RDT OR "point-of-care" OR POC) | Publication year: 2010 - 2025 |
| ClinicalTrials.gov | 12th January 2026 | (HIV) AND (diagnosis OR screening OR early HIV detection) AND (rapid diagnostic test OR RDT OR point-of-care OR POC OR test OR algorithm) AND (fourth-generation OR 4th | First Posted: 01.01.2010 - 31.12.2025 |

| | | | |
|---|---|---|---|
| | | generation OR Ag/Ab OR antigen-antibody OR Determine OR Alere) AND (acute HIV infection OR AHI OR early OR recent OR AEHI) | |
| Cochrane Library | 12th January 2026 | HIV AND ("diagnostic accuracy" OR sensitivity) AND (assay OR test OR "rapid diagnostic test" OR fourth generation OR Ag Ab OR antigen antibody OR "Alere HIV Combo" OR "point of care" OR "Determine HIV 1 2 Ag Ab Combo" OR "Determine Early Detect") | Publication year: 01.01.2010 - 31.12.2025 |
| Scopus | 12th January 2026 | TITLE-ABS-KEY(HIV OR "acute HIV" OR "early HIV") AND TITLE-ABS-KEY(diagnosis OR screening) AND TITLE-ABS-KEY(sensitivity OR "diagnostic accuracy" OR "diagnostic performance" OR detection) AND TITLE-ABS-KEY("fourth generation" OR "4th generation" OR Ag Ab OR antigen antibody) AND TITLE-ABS-KEY(test OR assay OR "rapid diagnostic test" OR RDT OR "point of care") AND PUBYEAR > 2009 AND PUBYEAR < 2026 | Publication year 2010 - 2025 adding to search string: PUBYEAR > 2009 AND PUBYEAR < 2026 |
| Web of Science | 12th January 2026 | ((((((AB=(Acute HIV)) OR AB=(HIV)) OR AB=(early HIV)) AND AB=(diagnosis OR screening)) AND AB=(sensitivity specificity OR diagnostic accuracy OR diagnostic performance OR detection)) AND AB=(fourth generation OR 4th generation OR Ag Ab OR antigen antibody)) AND AB=(test OR assay OR rapid diagnostic test OR RDT or point of care OR POC) | Publication year: 2006* - 2025 |

**Following the methodology from the WHO preprint study, the initial filter specified for this search was 2006. Any publications published before 2010 were filtered out in the PRISMA analysis.*

**Appendix S2. Table showing WHO prequalified fourth-generation HIV 1/2 Ag/Ab RDTs, approved as of 31 December 2025.**

| Product Name | WHO Product ID | Assay Format | Regulatory Version | Manu-facturer Name | Pathogen / Disease Marker | Year of Pre-qualification | Specimen Type | IVD Manufacturing Site(s) |
|---|---|---|---|---|---|---|---|---|
| Determine HIV Early Detect | 0243-013-00 | Immuno-chromatographic (lateral flow) | Rest-of-World | Abbott Diagnostics Medical Co., Ltd. | HIV | 2016 | Plasma, Serum, Whole blood (capillary), Whole blood (venous) | Abbott Diagnostics Medical Co., Ltd. (Formely "Alere Medical Co. Ltd.") 357 Matsuhidai, Chiba-ken Matsudoshi, 270-2214 Japan |

**Appendix S3. Risk of bias assessment using QUADAS-2.**

| First author, year of publication and Ag/Ab RDT evaluated. | Patient Selection | Index test | Reference standard | Flow and timing |
|---|---|---|---|---|
| Beelaert et al. 2010 | High | Unclear | Low | Low |
| Fox et al. 2011 | High | Unclear | Low | Low |
| Naylor et al. 2011 | Low | Low | Low | Low |
| Rosenberg et al. 2011 | Low | Low | Low | Low |
| Kilembe et al. 2012 | High | Unclear | Low | Low |
| Laperche et al. 2012 | High | High | Low | Low |
| Patel et al. 2012 | High | Low | Low | Low |
| Brauer et al. 2013 | High | Unclear | Low | Low |
| Faraoni et al. 2013 | High | Unclear | Low | Low |
| Kawahata et al. 2013 | High | Unclear | Low | Low |
| Pilcher et al. 2013 | High | High | Low | Low |
| Conway et al. 2014 | Low | Low | Low | Low |
| Duong et al. 2014 | Low | Low | Low | Low |
| Ottiger and Huber 2015 | High | Unclear | Low | Low |
| Smit et al. 2016 | High | High | Low | Low |
| Stekeler et al. 2016 | Low | Low | Low | Low |
| Delaugerre et al. 2017 | High | Unclear | Low | Low |
| Fitzgerald et al. 2017 | High | Unclear | Low | Low |
| Fransen et al. 2017 | High | Unclear | Low | Low |
| Livant et al. 2017 | High | Unclear | Low | Low |
| Masciotra et al. 2017 | Low | Low | Low | Low |
| Stafylis et al. 2017 | High | Unclear | Low | Low |
| Parker et al. 2018 | High | Unclear | Low | Low |
| Van Tienen et al. 2018 | High | High | Low | Low |
| Chavez et al. 2020 | Low | Low | Low | Low |
| Wratil et al. 2020 | High | Unclear | Low | Low |
| Kerschberger et al. 2021 | Low | Low | Low | Low |
| Alice Manjate et al. 2024 | Low | Low | Low | Low |
| Guiraud et al. 2024 | High | High | Low | Low |
| Ciglenecki et al. 2025 | Low | Low | Low | Low |
| Sirivichayakul et al. 2025 | High | Unclear | Low | Low |

#### Appendix S4. Overall sensitivity Ag/Ab.

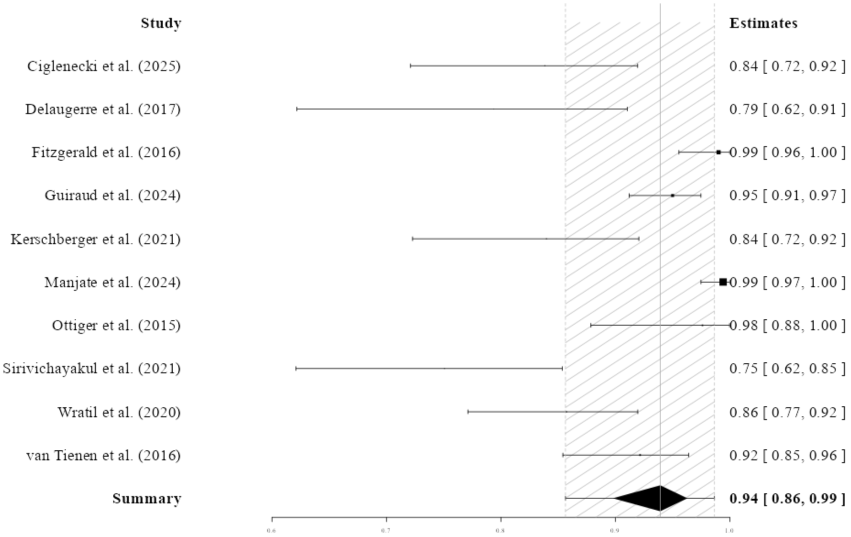

**Appendix S5. p24 Ag sensitivity in risk populations.**

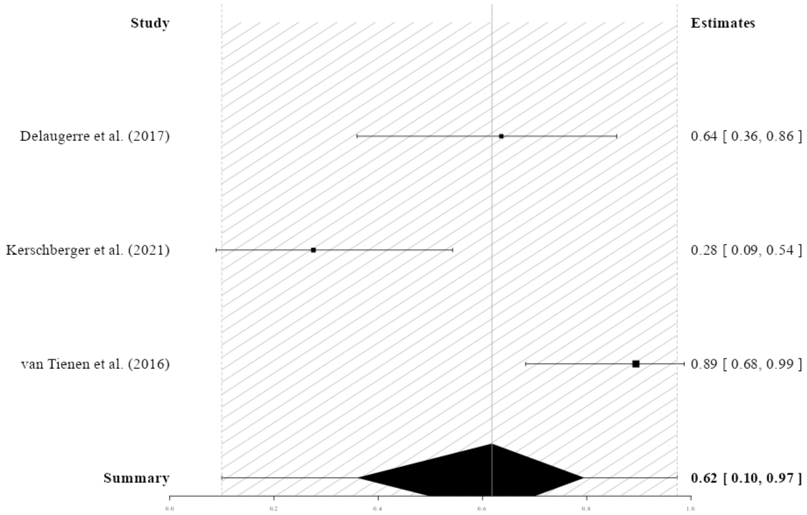

### Appendix S6. p24 Ag sensitivity in plasma/serum samples.

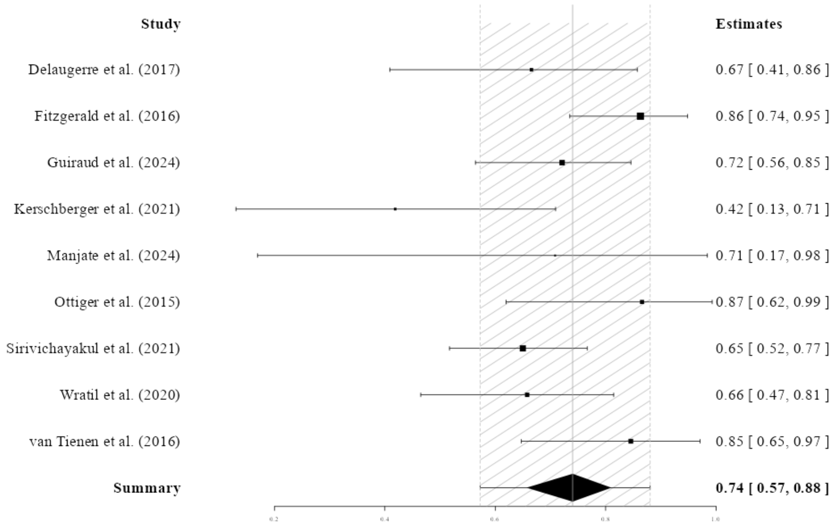
